# Persistent Presence of Spike protein and Viral RNA in the Circulation of Individuals with Post-Acute Sequelae of COVID-19

**DOI:** 10.1101/2022.08.07.22278520

**Authors:** Vaughn Craddock, Aatish Mahajan, Balaji Krishnamachary, Leslie Spikes, Prabhakar Chalise, Navneet K. Dhillon

## Abstract

SARS-CoV-2, the causative agent of COVID-19 disease has resulted in the death of millions worldwide since the beginning of the pandemic in December 2019. While much progress has been made to understand acute manifestations of SARS-CoV-2 infection, less is known about post-acute sequelae of COVID-19 (PASC). We investigated the levels of circulating SARS-CoV-2 components, Spike protein and viral RNA, in patients hospitalized with acute COVID-19 and in patients with and without PASC. In hospitalized patients with acute COVID-19 (n=116), we observed a positive correlation of Spike protein with D-dimer, length of hospitalization, and peak WHO score while viral RNA correlated with a tissue injury marker, lactate dehydrogenase (LDH). When comparing patients with post-COVID symptoms (n=33) and patients without (n=14), we found that Spike protein and viral RNA were more likely to be present in patients with PASC and in some cases at higher levels compared to acute COVID-19 patients. We also observed that the percent positivity of circulating viral RNA increased in the PASC positive individuals compared to acute COVID-19 group while Spike protein positivity remained the same. Additionally, we report that part of the circulating Spike protein is linked to extracellular vesicles without any presence of viral RNA in these vesicles. In conclusion, our findings suggest that Spike protein and/or viral RNA fragments persist in the recovered COVID-19 patients with PASC, regardless of their presence or absence during the acute COVID-19 phase. These results may help in understanding the reason(s) for why patients experience PASC symptoms and may offer potential therapeutic targets.

## INTRODUCTION

The COVID-19 pandemic caused by SARS-CoV-2 continues to have devastating global consequences with more than 579 million infections and over 6.4 million deaths internationally at the time of this writing^1^. The infection can progress to a severe illness that necessitates hospitalization and intensive care or can be mild and even asymptomatic. As the pandemic progressed, it quickly became apparent that a plethora of symptoms can persist long after the acute infection, no matter the initial severity. This has become known as Post-acute Sequelae of COVID-19 or PASC. Studies to date estimate that 30-50% of people who had COVID-19 still experience at least one post-COVID symptom 4 weeks after infection and 10-30% even after 12 weeks of acute infection^2, 3^. While the definition and scope of PASC remains to be determined, it is currently defined as new or persistent symptoms of COVID-19 beyond 4 weeks of acute infection that cannot be explained by other underlying etiologies^4, 5^. With the huge toll of acute infections that have and continue to occur worldwide, even if only a small fraction of patients goes on to develop PASC, it will have an enormous impact on society and the healthcare system for years to come. Therefore, it is imperative to understand the mechanism(s) of acute and post-acute COVID-19 disease pathogenesis, identify those most at risk of developing PASC, and investigate targets for future treatments.

Recent studies suggest that SARS-CoV-2 RNA can disseminate to extra-pulmonary tissues even in individuals with asymptomatic infection or mild COVID-19 disease and that viral RNA can reside in tissues for over 7 months in some individuals^6^. Multiple findings report SARS-CoV-2 Spike protein mediated lung injury and vascular damage by inducing endothelial barrier dysfunction and inflammation^7-9^. We previously reported significant apoptosis of microvascular endothelial cells on exposure to plasma-derived extracellular vesicles (EVs) from hospitalized COVID-19 patients^10^. Additionally, EVs released by lung epithelial cells transduced with lentivirus encoding SARS-CoV-2 proteins have been shown to transfer viral RNA to recipient cardiomyocytes, which might contribute to COVID-19-related myocardial inflammation and dysfunction^11^. Furthermore, some of the prolonged symptoms of PASC, such as persistent dyspnea, chest pain, and fatigue, suggest that the endothelial injury and thrombo-inflammation that occur in acute SARS CoV-2 infection may not resolve, thereby contributing to the persistence of symptoms^12^. A recent study reported the presence of microclots in the plasma from COVID-19 patients with PASC symptoms^13^. Therefore, in this study we examined if circulation of SARS-CoV-2 RNA or Spike protein is associated with acute disease severity and if its persistence correlated with manifestations of PASC. We report that both viral RNA and/or Spike protein remain in circulation long after acute infection (more than one-year post-infection in some cases) and this persistent circulation of viral components is associated with PASC. Further, we show the presence of Spike protein, but not viral RNA, in plasma-derived small EVs from individuals with acute or long-COVID-19.

## METHODS

### Sample and data collection

In this study we analyzed SARS-CoV-2 RNA and Spike protein in the plasma from a total of 151 patients either acutely infected and/or those who had recovered from acute infection.

#### Acute COVID-19

EDTA plasma samples (n=116) were obtained from patients admitted to the University of Kansas Health System (TUKHS) with a confirmed positive COVID-19 PCR test. The symptomatic, hospitalized COVID-19 patients (Acute) were categorized into three groups based on WHO Clinical Progression Scale score during hospitalization as mentioned in our previous publication^10, 14^ : 1) mild/moderate disease with no oxygen requirement (*Moderate-No O*_*2*,_ n=39), 2) moderate disease requiring supplemental oxygen (*Moderate-On O*_*2*,_ n*=*40), 3) severe disease (*Severe=37)*. Biospecimens in the Acute COVID biorepository were collected on the initial day of enrollment and every 3 days subsequently until discharge or death.

#### Post-acute sequelae of COVID-19 (PASC)

PASC patients were recruited when being seen in our Long-COVID clinic at TUKHS (N=33). This group was defined as having one or more persistent symptoms greater than 8 weeks post-infection, whether reported via a REDCAP survey or if they were being seen by a provider for persistent symptoms. These symptoms included fatigue, post-exertional malaise, shortness of breath, cough, loss of taste/smell, fever, myalgias, headaches, chest pain, “brain fog”, sleep disturbances, mood changes, rash, or tinnitus. Biospecimens were obtained at initial enrollment at least 18 weeks post-acute infection. PASC negative patients were identified through the ACTIV-2 trial participants after they had completed the first 24 weeks of the trial and defined as having a prior COVID-19 infection (PCR positive) greater than 8-12 weeks and did not have any persistent symptoms (N=14). We were also able to enroll 12 patients that were included in the Acute COVID Biorepository and follow them longitudinally in our PASC Biorepository.

These studies were approved by the local IRB at the University of Kansas Medical Center and all individuals gave consent. Demographic information, comorbid conditions, BMI, days between last positive test and study enrollment, and symptoms were all collected from acute and post-acute patients via electronic medical record or survey data and stored in a secure database.

Surveys were conducted through REDCap utilizing the standardized World Health Organization (WHO) established Global COVID-19 Clinical Platform Case Report Form (CRF) for Post-COVID conditions (Post COVID-19 CRF). Surveys were conducted at initial enrollment and subsequently every 3 months.

### Measurement of SARS-CoV-2 RNA Using Droplet Digital PCR

RNA was isolated using 200μl plasma as previously reported by others^15^. After centrifugation of plasma at approximately 21,000 × g for 2h at 4 °C, supernatant was removed and 750 μL of TRIzol-LS™ Reagent (ThermoFisher) was added to the pellet and incubated on ice for 10 minutes. Following incubation, 200 μL of chloroform (Millipore-Sigma) was added, vortexed and centrifuged at 21,000 × g for 15 min at 4 °C. Subsequently, the aqueous layer was removed and treated with an equal volume of isopropanol (Sigma) followed by addition of GlycoBlue™ Coprecipitant (ThermoFisher) and 100 μL 3 M Sodium Acetate (Life Technologies) and incubation on dry ice until frozen. RNA was later pelleted by centrifugation at 21,000 × g for 45 mins at 4 °C. The supernatant was then discarded, and the RNA pellet washed with cold 70% ethanol followed by resuspension in diethyl pyrocarbonate-treated water (ThermoFisher). SARS-CoV-2 RNA copies were quantified using the QX200TM Droplet Digital TM PCR System (ddPCR) and BioRad SARS-CoV-2 ddPCR Kit according to the manufacturer’s instruction. Data was analyzed using the QuantaSoftTM 1.7.4 Software (Bio-Rad) and SARS-CoV-2 quantification was expressed in number copies/μl. Also, to check the presence of free viral RNA, plasma was subjected to RNase A (0.5 μg/μl) treatment at 37°C for 20 minutes. Samples were then immediately subjected to RNA isolation followed by dd-PCR.

### Isolation of Extracellular Vesicles

Extracellular vesicles (EVs) were isolated from EDTA plasma using Ultracentrifugation. Hemolysis of blood was ruled out by visually inspecting plasma samples for any pink discoloration indicative of hemolysis or by measuring absorbance of hemoglobin at 414nm. About 1ml of platelet-free plasma (PFP) was centrifuged at 20,000 x g for 15 minutes at 4ºC to remove large sized EVs. The supernatants were then centrifuged at 100,000 x g for 70 minutes at 4ºC, washed with PBS, and spun again at 100,000 x g for 70 minutes to isolate small EVs *(SEVs*). The abundance and size distribution and purity of small EVs was compared among different groups using the NanoSight LM10 system, TEM and Western blot analysis of EV markers as mentioned in our previous publication^10^.

### Spike protein ELISA

The levels of SARS-CoV-2 Spike protein in the plasma and EV samples were determined using the SAR-CoV-2 (2019-nCoV) Spike protein ELISA Kit according to the manufacturer instructions (SinoBiological). To check the levels of Spike protein in EVs treated with proteinase K, 100μg of Spike protein positive COVID-19 EVs were treated with and without 50 μg/ml of proteinase K (Exiqon) for 30 min at 37 °C. Then the reaction was inhibited by incubation with 5mM phenylmethylsulfonyl fluoride (PMSF) for 10 min at 37 °C (Mol. Pharmaceutics 2018, 15, 3, 1073–1080). EVs were also treated with and without heparinase II (0.9 mIU/mL, New England Biolabs) for 3 h at 37 °C and then incubated again with fresh enzyme overnight at 37°C ^16^. After proteinase K or heparinase treatment, EVs were washed with PBS and concentrated using Amicon Ultra-4 centrifugal filters (10 kDa, Millipore Sigma, USA).

### Statistical Analysis

The demographic and clinical characteristics for the three disease severity groups (*Moderate-No O*_*2*_, *Moderate-On O*_*2*_ and *Severe*) were summarized with median and interquartile range for the continuous variables and with frequencies and percentages for categorical variables, (Table 1). The differences in the continuous variables among the groups were assessed using Kruskal Wallis Test. The categorical outcomes among the groups were assessed using Fisher exact test. Spearman’s rank correlation analyses were carried out to assess the relationship between each of the plasma Spike protein, EV-Spike protein and viral RNA with Age, BMI, WBC, lymphocyte count, CRP, creatinine, D-dimer etc. Similar analyses were carried out for the outcomes between positive and negative status of PASC (Table 2). The differences in the continuous variables between PASC-negative and PASC-positive groups were assessed using Wilcoxon rank sum test. Categorical variables were assessed using Fisher Exact test and Spearman’s rank correlation analysis of each of Spike protein and viral RNA were carried out with Age, BMI, and days of hospitalization during acute infection. All the tests were considered statistically significant if p-values were less than 0.05. The statistical analyses were carried out using the statistical software R version 4.0.0 (R core Team, 2020).

**Table 1:**
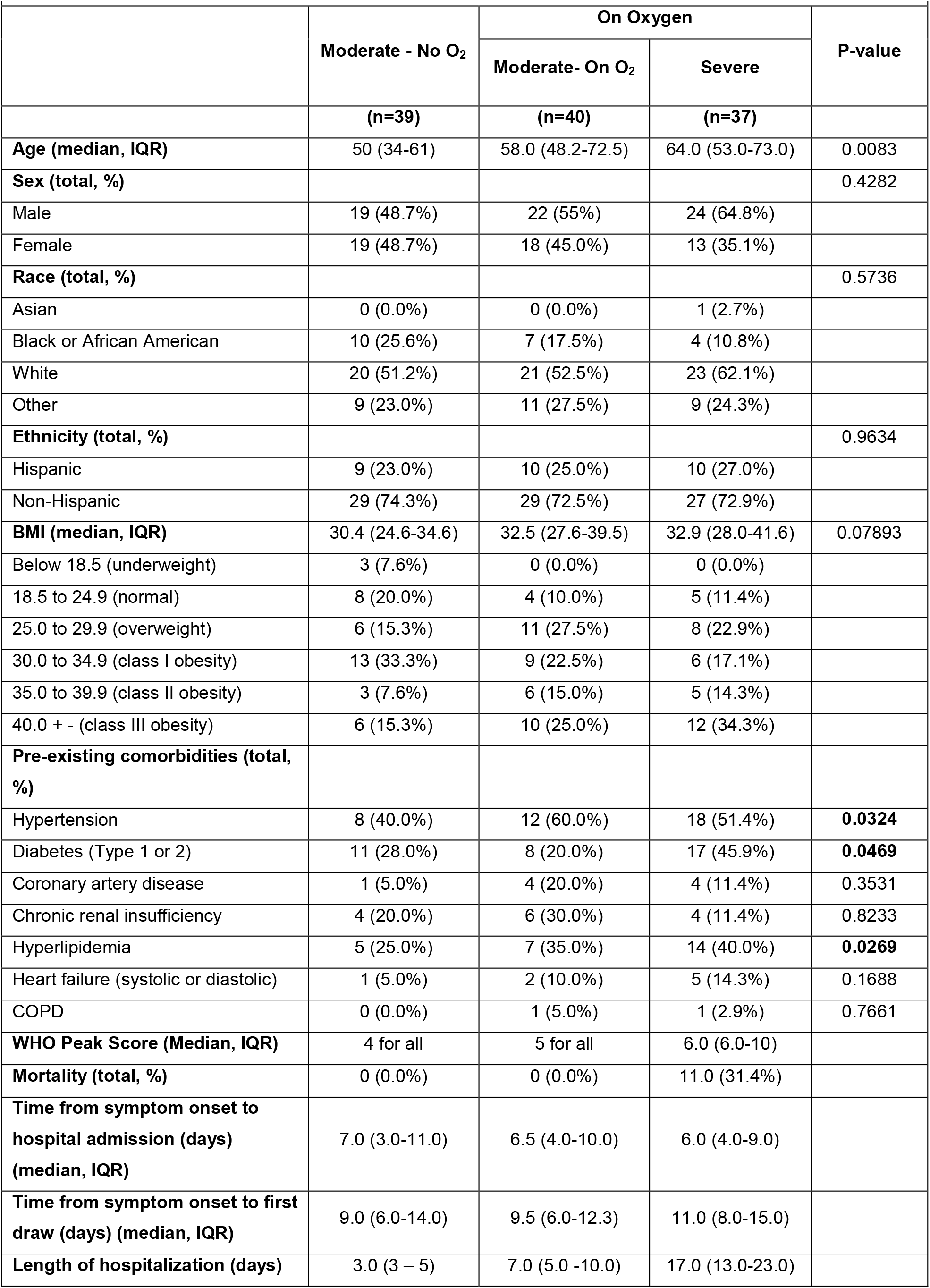
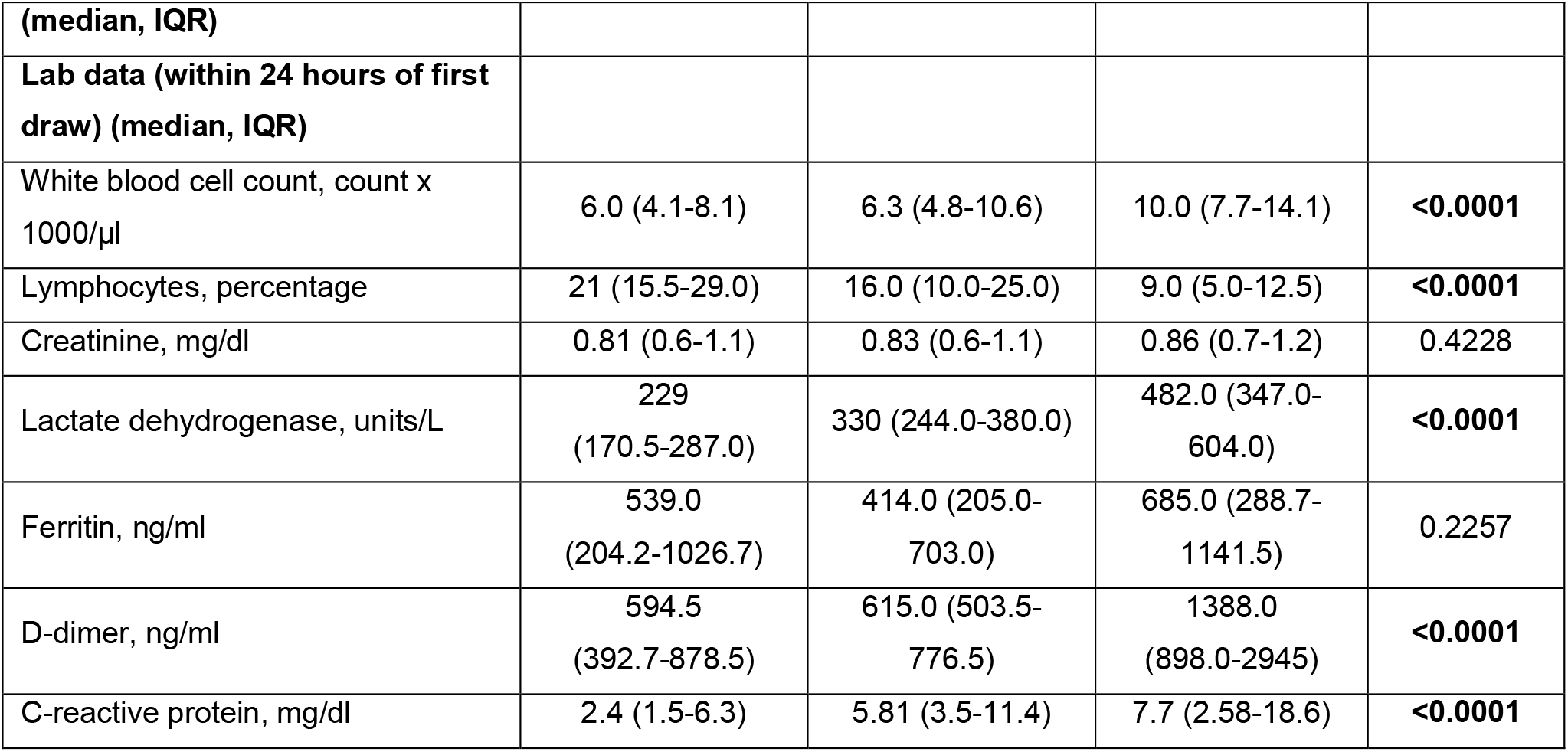
Demographics & Characteristics of Acute COVID-19 Patients Included in Analysis

|  | Moderate - No O<sub>2</sub> | On Oxygen |  | P-value |
| --- | --- | --- | --- | --- |
|  |  | Moderate- On O<sub>2</sub> | Severe |  |
|  | (n=39) | (n=40) | (n=37) |  |
| **Age (median, IQR)** | 50 (34-61) | 58.0 (48.2-72.5) | 64.0 (53.0-73.0) | 0.0083 |
| **Sex (total, %)** |  |  |  | 0.4282 |
| Male | 19 (48.7%) | 22 (55%) | 24 (64.8%) |  |
| Female | 19 (48.7%) | 18 (45.0%) | 13 (35.1%) |  |
| **Race (total, %)** |  |  |  | 0.5736 |
| Asian | 0 (0.0%) | 0 (0.0%) | 1 (2.7%) |  |
| Black or African American | 10 (25.6%) | 7 (17.5%) | 4 (10.8%) |  |
| White | 20 (51.2%) | 21 (52.5%) | 23 (62.1%) |  |
| Other | 9 (23.0%) | 11 (27.5%) | 9 (24.3%) |  |
| **Ethnicity (total, %)** |  |  |  | 0.9634 |
| Hispanic | 9 (23.0%) | 10 (25.0%) | 10 (27.0%) |  |
| Non-Hispanic | 29 (74.3%) | 29 (72.5%) | 27 (72.9%) |  |
| **BMI (median, IQR)** | 30.4 (24.6-34.6) | 32.5 (27.6-39.5) | 32.9 (28.0-41.6) | 0.07893 |
| Below 18.5 (underweight) | 3 (7.6%) | 0 (0.0%) | 0 (0.0%) |  |
| 18.5 to 24.9 (normal) | 8 (20.0%) | 4 (10.0%) | 5 (11.4%) |  |
| 25.0 to 29.9 (overweight) | 6 (15.3%) | 11 (27.5%) | 8 (22.9%) |  |
| 30.0 to 34.9 (class I obesity) | 13 (33.3%) | 9 (22.5%) | 6 (17.1%) |  |
| 35.0 to 39.9 (class II obesity) | 3 (7.6%) | 6 (15.0%) | 5 (14.3%) |  |
| 40.0 + - (class III obesity) | 6 (15.3%) | 10 (25.0%) | 12 (34.3%) |  |
| **Pre-existing comorbidities (total, %)** |  |  |  |  |
| Hypertension | 8 (40.0%) | 12 (60.0%) | 18 (51.4%) | **0.0324** |
| Diabetes (Type 1 or 2) | 11 (28.0%) | 8 (20.0%) | 17 (45.9%) | **0.0469** |
| Coronary artery disease | 1 (5.0%) | 4 (20.0%) | 4 (11.4%) | 0.3531 |
| Chronic renal insufficiency | 4 (20.0%) | 6 (30.0%) | 4 (11.4%) | 0.8233 |
| Hyperlipidemia | 5 (25.0%) | 7 (35.0%) | 14 (40.0%) | **0.0269** |
| Heart failure (systolic or diastolic) | 1 (5.0%) | 2 (10.0%) | 5 (14.3%) | 0.1688 |
| COPD | 0 (0.0%) | 1 (5.0%) | 1 (2.9%) | 0.7661 |
| **WHO Peak Score (Median, IQR)** | 4 for all | 5 for all | 6.0 (6.0-10) |  |
| **Mortality (total, %)** | 0 (0.0%) | 0 (0.0%) | 11.0 (31.4%) |  |
| **Time from symptom onset to hospital admission (days) (median, IQR)** | 7.0 (3.0-11.0) | 6.5 (4.0-10.0) | 6.0 (4.0-9.0) |  |
| **Time from symptom onset to first draw (days) (median, IQR)** | 9.0 (6.0-14.0) | 9.5 (6.0-12.3) | 11.0 (8.0-15.0) |  |
| **Length of hospitalization (days)** | 3.0 (3 – 5) | 7.0 (5.0 -10.0) | 17.0 (13.0-23.0) |  |
| (median, IQR) |  |  |  |  |
| Lab data (within 24 hours of first draw) (median, IQR) |  |  |  |  |
| White blood cell count, count x 1000/μl | 6.0 (4.1-8.1) | 6.3 (4.8-10.6) | 10.0 (7.7-14.1) | **<0.0001** |
| Lymphocytes, percentage | 21 (15.5-29.0) | 16.0 (10.0-25.0) | 9.0 (5.0-12.5) | **<0.0001** |
| Creatinine, mg/dl | 0.81 (0.6-1.1) | 0.83 (0.6-1.1) | 0.86 (0.7-1.2) | 0.4228 |
| Lactate dehydrogenase, units/L | 229<br>(170.5-287.0) | 330 (244.0-380.0) | 482.0 (347.0-604.0) | **<0.0001** |
| Ferritin, ng/ml | 539.0<br>(204.2-1026.7) | 414.0 (205.0-703.0) | 685.0 (288.7-1141.5) | 0.2257 |
| D-dimer, ng/ml | 594.5<br>(392.7-878.5) | 615.0 (503.5-776.5) | 1388.0<br>(898.0-2945) | **<0.0001** |
| C-reactive protein, mg/dl | 2.4 (1.5-6.3) | 5.81 (3.5-11.4) | 7.7 (2.58-18.6) | **<0.0001** |

**Table 2:**
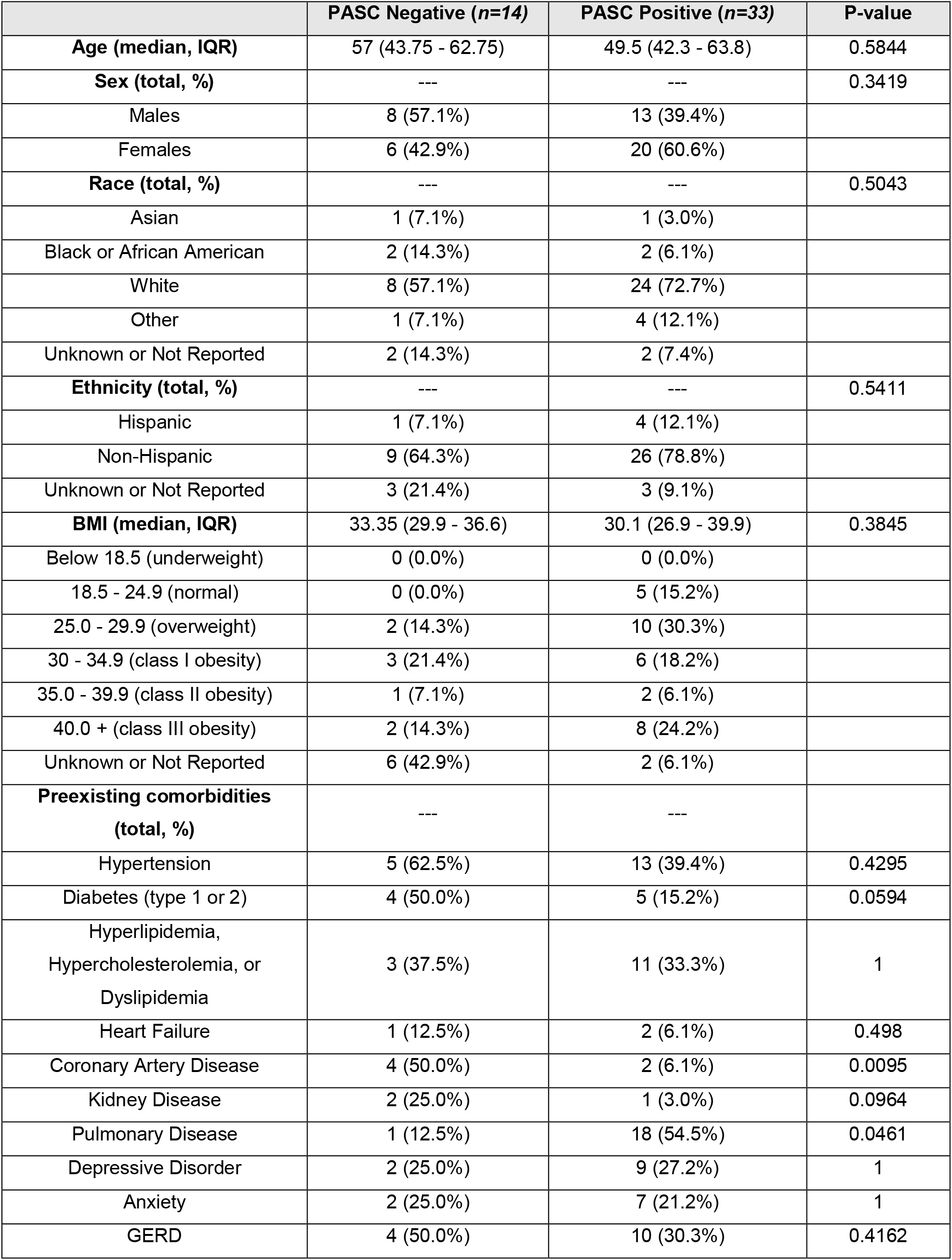
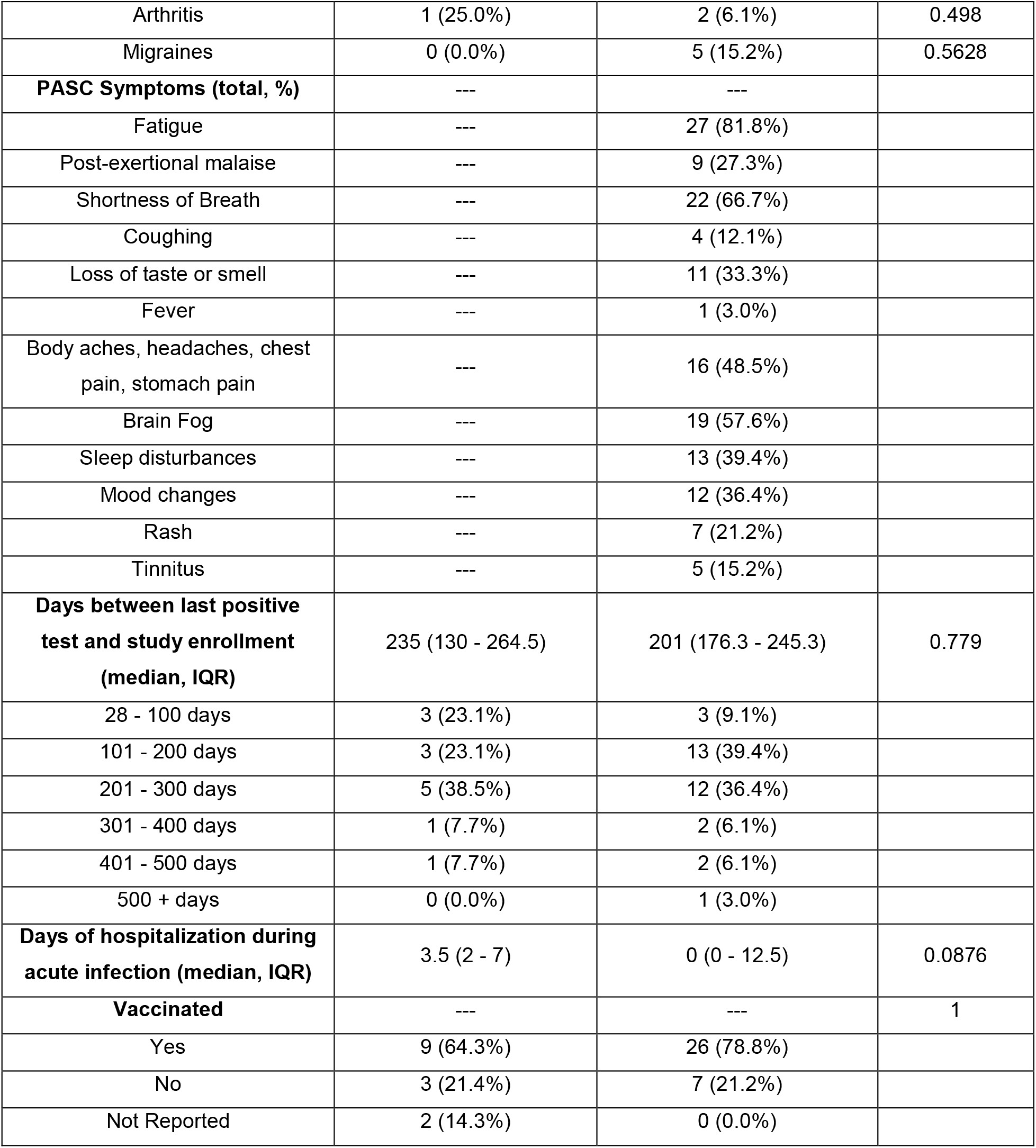
Demographics & Characteristics of PASC Patients Included in Analysis

|  |  |  |  |
| --- | --- | --- | --- |
|  | PASC Negative (n=14) | PASC Positive (n=33) | P-value |
| **Age (median, IQR)** | 57 (43.75 - 62.75) | 49.5 (42.3 - 63.8) | 0.5844 |
| **Sex (total, %)** | --- | --- | 0.3419 |
| Males | 8 (57.1%) | 13 (39.4%) |  |
| Females | 6 (42.9%) | 20 (60.6%) |  |
| **Race (total, %)** | --- | --- | 0.5043 |
| Asian | 1 (7.1%) | 1 (3.0%) |  |
| Black or African American | 2 (14.3%) | 2 (6.1%) |  |
| White | 8 (57.1%) | 24 (72.7%) |  |
| Other | 1 (7.1%) | 4 (12.1%) |  |
| Unknown or Not Reported | 2 (14.3%) | 2 (7.4%) |  |
| **Ethnicity (total, %)** | --- | --- | 0.5411 |
| Hispanic | 1 (7.1%) | 4 (12.1%) |  |
| Non-Hispanic | 9 (64.3%) | 26 (78.8%) |  |
| Unknown or Not Reported | 3 (21.4%) | 3 (9.1%) |  |
| **BMI (median, IQR)** | 33.35 (29.9 - 36.6) | 30.1 (26.9 - 39.9) | 0.3845 |
| Below 18.5 (underweight) | 0 (0.0%) | 0 (0.0%) |  |
| 18.5 - 24.9 (normal) | 0 (0.0%) | 5 (15.2%) |  |
| 25.0 - 29.9 (overweight) | 2 (14.3%) | 10 (30.3%) |  |
| 30 - 34.9 (class I obesity) | 3 (21.4%) | 6 (18.2%) |  |
| 35.0 - 39.9 (class II obesity) | 1 (7.1%) | 2 (6.1%) |  |
| 40.0 + (class III obesity) | 2 (14.3%) | 8 (24.2%) |  |
| Unknown or Not Reported | 6 (42.9%) | 2 (6.1%) |  |
| **Preexisting comorbidities (total, %)** | --- | --- |  |
| Hypertension | 5 (62.5%) | 13 (39.4%) | 0.4295 |
| Diabetes (type 1 or 2) | 4 (50.0%) | 5 (15.2%) | 0.0594 |
| Hyperlipidemia, Hypercholesterolemia, or Dyslipidemia | 3 (37.5%) | 11 (33.3%) | 1 |
| Heart Failure | 1 (12.5%) | 2 (6.1%) | 0.498 |
| Coronary Artery Disease | 4 (50.0%) | 2 (6.1%) | 0.0095 |
| Kidney Disease | 2 (25.0%) | 1 (3.0%) | 0.0964 |
| Pulmonary Disease | 1 (12.5%) | 18 (54.5%) | 0.0461 |
| Depressive Disorder | 2 (25.0%) | 9 (27.2%) | 1 |
| Anxiety | 2 (25.0%) | 7 (21.2%) | 1 |
| GERD | 4 (50.0%) | 10 (30.3%) | 0.4162 |
| Arthritis | 1 (25.0%) | 2 (6.1%) | 0.498 |
| Migraines | 0 (0.0%) | 5 (15.2%) | 0.5628 |
| **PASC Symptoms (total, %)** | --- | --- |  |
| Fatigue | --- | 27 (81.8%) |  |
| Post-exertional malaise | --- | 9 (27.3%) |  |
| Shortness of Breath | --- | 22 (66.7%) |  |
| Coughing | --- | 4 (12.1%) |  |
| Loss of taste or smell | --- | 11 (33.3%) |  |
| Fever | --- | 1 (3.0%) |  |
| Body aches, headaches, chest pain, stomach pain | --- | 16 (48.5%) |  |
| Brain Fog | --- | 19 (57.6%) |  |
| Sleep disturbances | --- | 13 (39.4%) |  |
| Mood changes | --- | 12 (36.4%) |  |
| Rash | --- | 7 (21.2%) |  |
| Tinnitus | --- | 5 (15.2%) |  |
| **Days between last positive test and study enrollment (median, IQR)** | 235 (130 - 264.5) | 201 (176.3 - 245.3) | 0.779 |
| 28 - 100 days | 3 (23.1%) | 3 (9.1%) |  |
| 101 - 200 days | 3 (23.1%) | 13 (39.4%) |  |
| 201 - 300 days | 5 (38.5%) | 12 (36.4%) |  |
| 301 - 400 days | 1 (7.7%) | 2 (6.1%) |  |
| 401 - 500 days | 1 (7.7%) | 2 (6.1%) |  |
| 500 + days | 0 (0.0%) | 1 (3.0%) |  |
| **Days of hospitalization during acute infection (median, IQR)** | 3.5 (2 - 7) | 0 (0 - 12.5) | 0.0876 |
| **Vaccinated** | --- | --- | 1 |
| Yes | 9 (64.3%) | 26 (78.8%) |  |
| No | 3 (21.4%) | 7 (21.2%) |  |
| Not Reported | 2 (14.3%) | 0 (0.0%) |  |

## RESULTS

### Presence of Viral RNA, soluble Spike protein and/or EV-linked Spike protein in the plasma of COVID-19 patients with acute disease

We compared hospitalized, symptomatic COVID-19 patients based on WHO disease severity score. As shown in Table 1, the age of patients was significantly different across the groups (p=0.008), with highest median age in the *Severe* group compared to others. There were significant differences in hypertension (p=0.0324), hyperlipidemia (p=0.0269), and diabetes (p=0.0469) as well other clinical lab parameters of inflammation, tissue injury and coagulopathy across groups. The reverse transcription-droplet digital polymerase chain reaction (RT-ddPCR) analysis revealed the presence of viral RNA in all three groups. We found viral RNA in the plasma from 33% of *Moderate-No O*_*2*_, 35% of *Moderate-On O*_*2*_ and 37% of *Severe* group patients without any significant differences in copy numbers across the groups (Figure 1A (i)). However, the levels of viral RNA in these acute COVID-19 patients showed significant positive correlation with tissue injury marker lactate dehydrogenase (LDH) (p=0.027). While we didn’t see overall correlation of viral RNA with BMI, viral RNA levels significantly correlated with BMI (p=0.009) in the *Severe* group patients. Further, to check if this positivity is due to the presence of free viral RNA in circulation, RNAse treatment was given to a small set of samples (n=10) before ddPCR analysis, As shown in Figure 1A (ii), viral RNA levels dropped in most of the samples, however some samples still showed high levels of viral RNA which could be due to existence of free virions in the plasma. Another possibility could be that the incubation time with RNAse was too short to completely degrade the free RNA in the samples with higher viral RNA levels to begin with.

**Figure 1:**
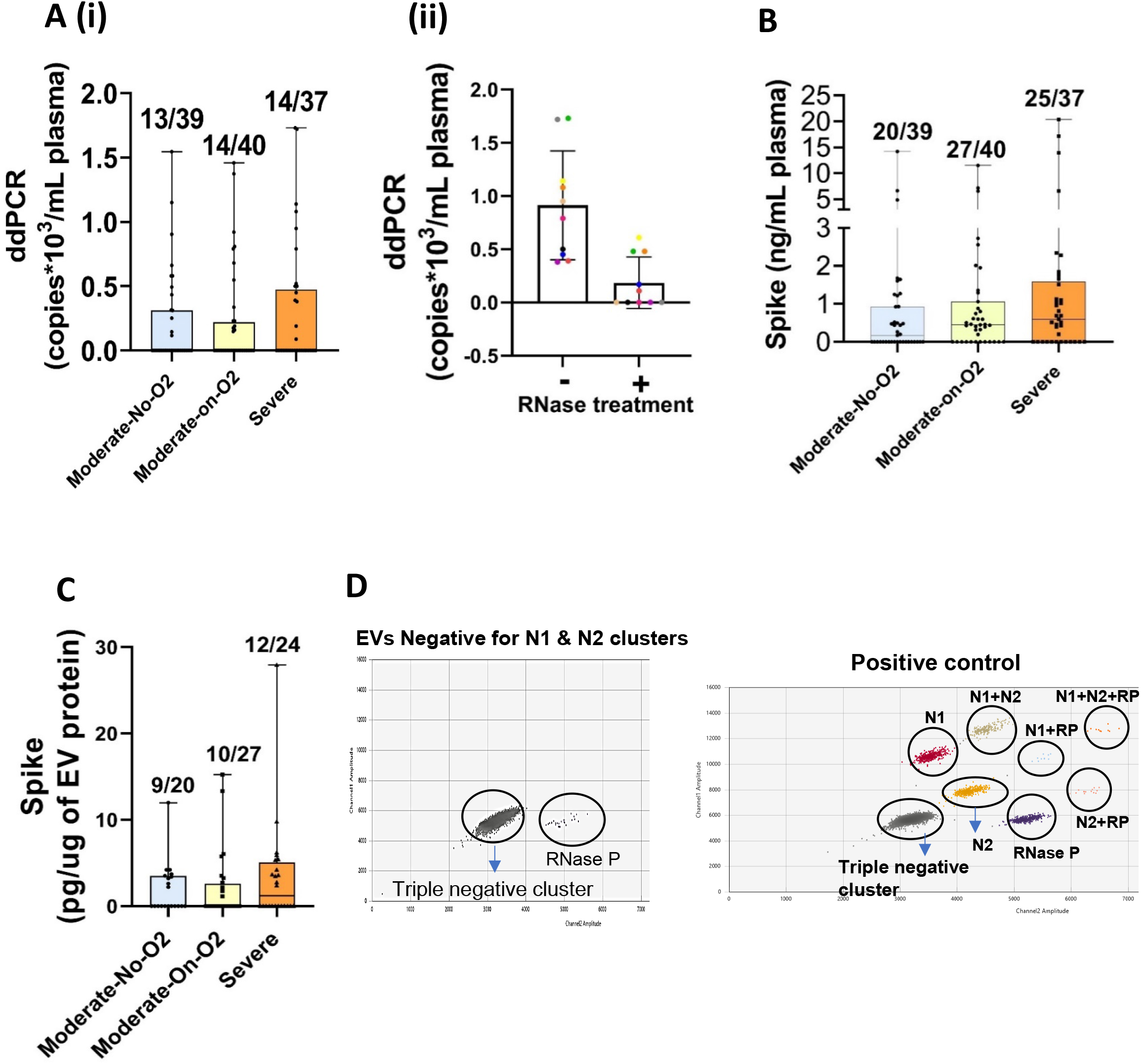
The presence of SARS-CoV-2 RNA and Spike protein in the EDTA-plasma of symptomatic hospitalized COVID-19 patients (Acute Disease). Patients (N-116) were grouped into three groups of disease severity based on WHO progression scale score: Moderate-No O2, (N=39), Moderate-On O2, (N=40) and Severe (N=37) groups. **A**) SARS-CoV-2 RNA copies in the plasma as measured by dd-PCR comparing all groups (Ai) and Viral RNA copies after RNase treatment (N=10) (Aii). **B**) Spike protein levels as detected by ELISA in the plasma from three groups. **C**) Spike ELISA in plasma derived small EVs from the subjects that showed positivity for Spike protein in total plasma. **D)** Representative 2D plot of dd-PCR showing absence of viral RNA in EVs as detected by the presence of internal control (RP) but absence of Nucleocapsid cluster. EVs from N=11 subjects that showed positivity for viral RNA and/or Spike protein in the corresponding plasma were analyzed. Also shown is the 2D plot of positive control (synthetic RNA transcript containing five gene targets: E, N, ORF1ab, RdRP and S Genes of SARS-CoV-2) showing the presence of nucleocapsid (N) cluster.

The analysis of plasma for Spike protein by ELISA showed the presence of Spike protein in 51% of *Moderate-No O*_2_, 68% of *Moderate-On O*_*2*_ and 68% of *Severe* group patients (Figure 1B). Despite the differences observed in comorbidities or age, we didn’t find significant differences in the Spike protein levels across the three groups. Nevertheless, the level of Spike protein showed significant positive correlation with peak WHO score (p=0.03) along with D-Dimer (p=0.02) and length of hospitalization (p=0.03) (Supplementary table 1). Overall, we found 35% of viral RNA positivity and 62% Spike protein positivity in symptomatic acute COVID-19 patients tested. Some of the samples that had circulating Spike protein did not have detectable levels of viral RNA, while other samples that had viral RNA did not have detectable Spike protein. While 34% of acute COVID-19 patients were positive for only Spike protein (30% *Moderate-No O*_2_, 40% *Moderate-On O*_*2*_ and 32% *Severe)*, positivity for viral RNA alone was observed in only 6% of total samples analyzed (10% *Moderate-No O*_2_, 5% *Moderate-On O*_*2*_ and 1% *Severe)*. This suggests that Spike protein may be freely circulating independent of circulating virions and circulating viral RNA may include free viral RNA or its fragments during acute infection with SARS-CoV-2. In parallel, to confirm the specificity of our viral RNA and Spike protein ELISA analyses, we examined viral RNA and Spike protein in the plasma samples from a cohort of patients never infected with COVID-19 as negative controls (Supplementary figure 1 A & B).

Next, the plasma samples which showed positivity for Spike protein were used to check for Spike protein in EVs. The results indicated that 45% of Spike protein positive *Moderate-No O*_*2*_ samples were positive for EV-linked Spike protein while 37% and 50% of *Moderate-on O*_*2*_ and *Severe* group individuals, respectively, showed presence of Spike protein in EVs (Figure 1C). The level of Spike protein-linked EVs significantly correlated with D-Dimer (p=0.009) in these acute group of COVID-19 patients (Supplementary Table 1). We also checked for the presence of viral RNA using ddPCR in EVs from the *Severe* group (n=11) that showed positivity for both Spike protein and viral RNA (n=8/11) and in those samples which showed very high levels of Spike protein but no viral RNA (n=3/11). Interestingly, we were not able to detect viral RNA in any of the EV samples tested (Figure 1D).

### Persistent circulation of Viral RNA and Spike protein is associated with PASC

We grouped PASC patients into PASC-positive (n=33) and PASC-negative (n=14) based on self-reported symptoms either from visits with a physician or questionnaires. Demographics and information on the PASC symptoms can be found in Table 2. The pre-existing comorbidities in these patients were diagnosed before patients developed PASC symptoms and the PASC symptoms they experienced were reported more than 28 days after their acute SARS-CoV-2 infection. Patients with PASC symptoms were more likely to be female (67%) but we did not see a statistically significant difference (p=0.3419). Patients with PASC were more likely to have pre-existing pulmonary disease (p=0.0461) which in most cases was mild-to-moderate asthma or obstructive sleep apnea. Patients without PASC symptoms (PASC-ve) were more likely to have diabetes (p=0.0594), coronary artery disease (p=0.0095), or kidney disease (p=0.0964). Interestingly, PASC-ve patients, including patients with diabetes, coronary artery disease, or kidney disease, were more likely to be hospitalized from acute COVID-19 (p=0.0876). Finally, there was no difference in vaccination status between PASC-positive (PASC+ve) and negative groups. We screened to ensure that administrations of mRNA vaccines were not confounding the levels of Spike protein detected in PASC patients. The Infectious Disease Society of America (IDSA) estimates that the Spike protein from mRNA vaccines is no longer detectable after a few weeks. The earliest time between a recent vaccination and blood draw for this study was 21 days in one patient, and this patient was PASC-ve and did not have detectable levels of Spike protein. All other patients in this study had their most recent vaccination more than 30 days before their consent and blood drawn for this study or received a non-mRNA vaccine.

When we examined the viral RNA copies in the plasma from PASC+ve and PASC-ve individuals, 28% of PASC-ve samples showed presence of viral RNA copies ranging from 0.08-2.13 copies/ul, whereas 59% of PASC+ve patients had presence of viral RNA in circulation (0.07-12.74 copies/ul) (Figure 2Ai) with much higher copy numbers in some of the individuals compared to PASC-ve or acute COVID-19 groups (Figure 1Ai). RNase treatment of plasma before RNA isolation resulted in the drop of viral RNA copies in most of the samples from the PASC+ve group (n=7) as illustrated in (Figure 2Aii). Furthermore, viral RNA in plasma showed a strong positive correlation with days of hospitalization (p=0.043). While circulating viral RNA had no correlation with BMI in PASC+ve group, a significant correlation was observed in PASC-ve samples (p=0.008) (Supplementary Table 1).

**Figure 2:**
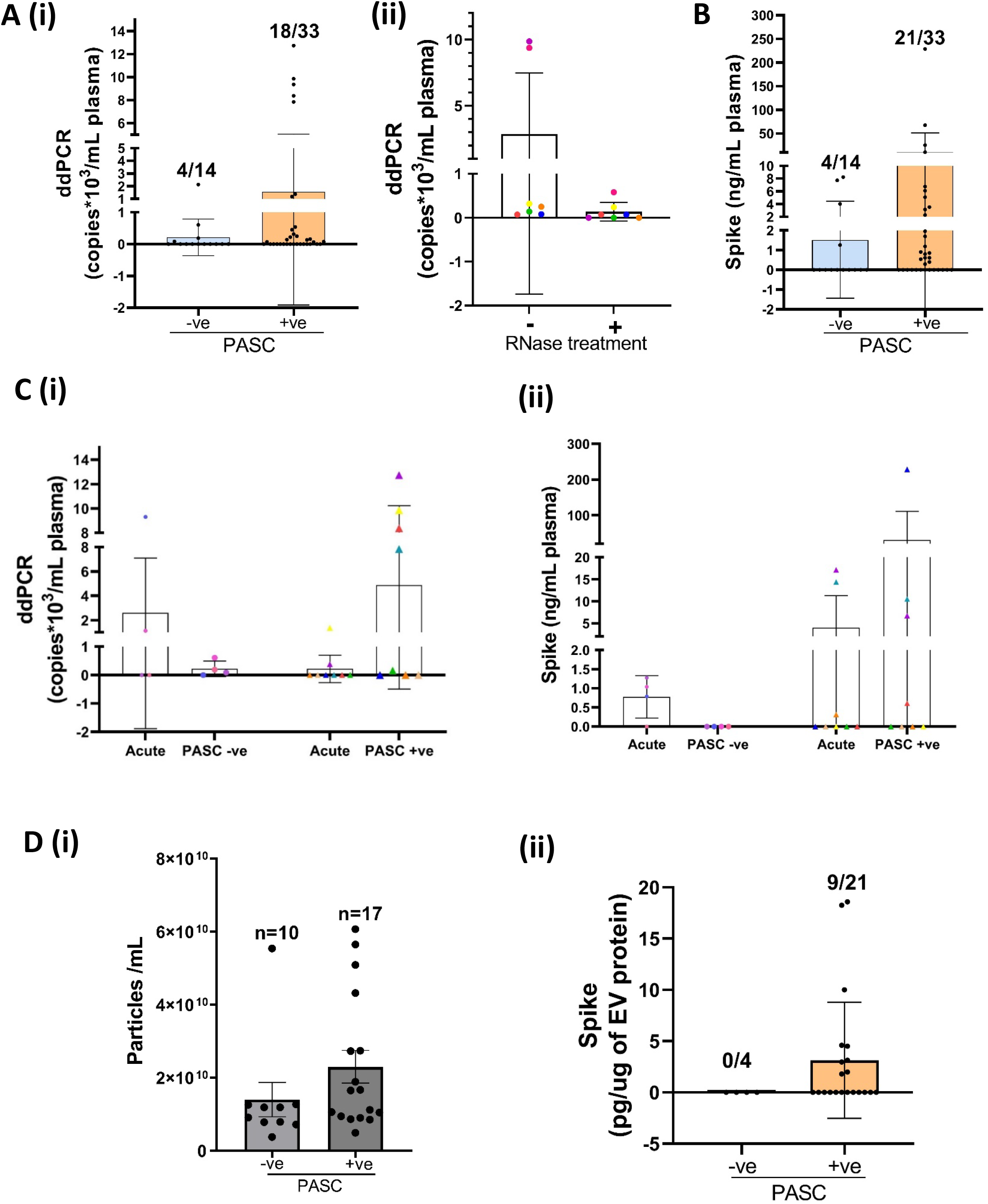
Viral RNA and/or Spike protein persistence in circulation is associated with PASC. Viral RNA (**A**) and Spike protein (**B**) levels were analyzed in the plasma from COVID -19 patients with PASC (persistent symptoms of COVID-19 at 8-12 weeks or more) (PASC +ve, N=33) and COVID-19 recovered patients that did not have any persistent symptoms (PASC-ve, N=14). SARS-CoV-2 RNA copies were measured using droplet digital -PCR and Spike protein by ELISA. **C**) Longitudinal analysis of viral RNA (Ci) and Spike protein (C ii) in individuals (N=12) at acute and post-acute phase of COVID-19 disease with and without PASC. **D**) (i) Comparison of small EV numbers (i) and EV-linked Spike protein (ii) in the plasma from individuals with and without PASC as measured by Nanoparticle Tracking Analysis and ELISA, respectively.

Parallel evaluation of Spike protein in the plasma also found the presence of Spike protein in 4 of 14 PASC-ve individuals and 21 of 33 of PASC+ve (64%) samples (Figure 2B) and the levels were within the range of the levels observed in acute COVID-19 samples. Additionally, we found that in PASC+ve group 33% (11/33) samples showed positivity for both Spike protein as well as viral RNA in plasma, 30% (10/33) for only Spike protein and 18% (6/33) for only viral RNA. However, in the PASC-ve group, none of the samples showed positivity for both Spike protein and viral RNA. When we compared the levels of viral RNA and Spike protein during the acute COVID-19 and post-recovery phase of COVID-19 in the same patients (n=12), we observed that in the patients from PASC+ve group, Spike protein and/or viral RNA increased or remained the same as in the acute phase; whereas, in the PASC-ve group, Spike protein was found to be totally absent and viral RNA either decreased to very low copy numbers or was undetectable (Figure 2 C).

Next, we isolated small EVs from the plasma of PASC-ve and PASC+ve samples that showed positivity for either Spike protein and/or viral RNA. Nanoparticle tracking analysis revealed a trend towards an increase in total number of EVs in the PASC+ve group as compared to the PASC-ve group (p=0.203) (Figure 2D,i). The lack of significant difference was potentially due to one of the PASC-ve individuals that showed a much higher number of EVs compared to rest of the subjects and had multiple underlying comorbidities, including diabetes, heart failure, and kidney disease. When we examined the Spike protein levels in the EVs from the plasma samples that showed positivity for Spike protein, 53% (8/15) of samples from the PASC+ve group showed presence of EV-linked Spike protein and these levels were almost at a similar level as that observed in acute COVID-19 patients (Figure 2D, ii). Interestingly, EVs from the PASC-ve group did not show any positivity for Spike protein ELISA.

### Presence of Spike protein on the surface of EVs

To test if Spike protein is on the surface of EVs, we first compared the levels of Spike protein in the EVs treated with or without proteinase K. To have enough protein for the analysis of Spike protein and to run western blots, we pooled two patient samples each for a set of n=3. As shown in Figure 3A, a significant decrease was observed in the levels of Spike protein in the EV preparation from ‘*Severe’* group patients after the proteinase K treatment. In parallel, we confirmed the removal of membrane proteins by proteinase K treatment by examining the levels of tetraspanins in EV preparations before and after proteinase K treatment. As expected proteinase K treatment resulted in the degradation of tetraspanins CD81 and CD9 from the EV membrane.

**Figure 3:**
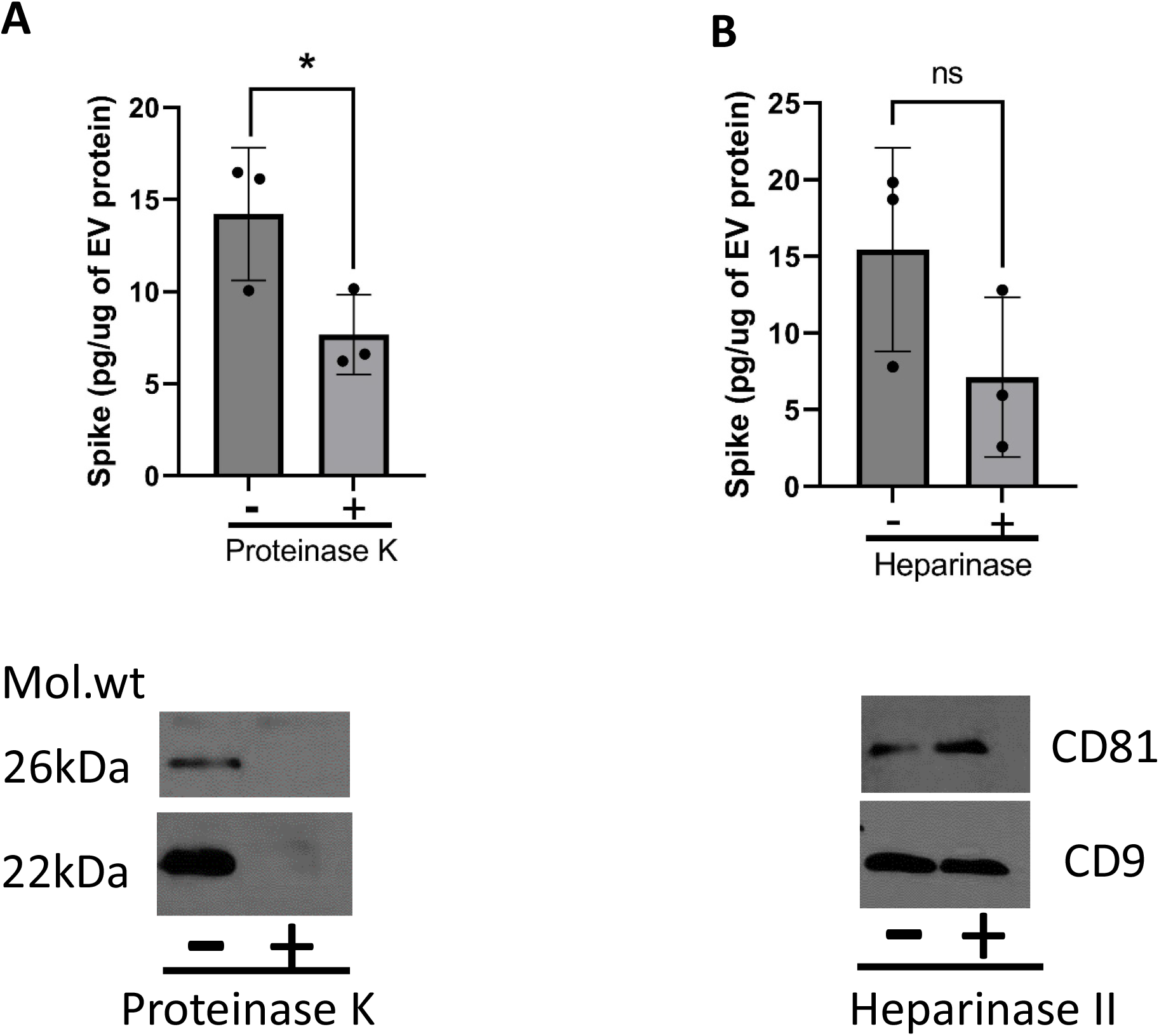
Spike protein is partly associated on the surface of EVs. Decrease in levels of Spike in EVs treated with proteinase K (**A**) and heparinase II (**B**) in comparison to untreated SEVs, as detected by ELISA. Western blots below showing loss of EV surface tetraspanins confirms the activity of proteinase K while the heparinase II treatment didn’t show any effect on these surface markers. EVs from 2 patients were pooled in a set of n=3 for the analysis. * p≤0.05

Cytokine and growth factors have been found to be linked to the EV surface through the heparan sulfate proteoglycans and Clausen et al. have reported that Spike protein interacts with heparan sulfate through the receptor binding domain^17^. Therefore, to assess if Spike protein is associated on EV surfaces by binding to heparan sulfate, we examined the levels of Spike protein in heparinase II-treated EVs. Heparinase II treatment of COVID-19 EVs showed decreased levels of Spike protein when compared to untreated EVs (Figure 3B, n=3, each a pool of EVs from 2 patients) where as CD81 and CD9 tetraspanins were still present on EVs after heparinase II treatment. This suggests specific interactions of Spike protein on EV surfaces.

## DISCUSSION

We report here that Spike protein and viral RNA circulate in acute COVID-19 patients, can persist for up to 1 year or longer after acute SARS-CoV-2 infection, and are associated with PASC. Our observations reveal that patients who have PASC were more likely to have higher levels of these SARS-CoV-2 components in circulation than those without PASC and that viral RNA was increased in PASC patients even in comparison to acutely infected individuals. Interestingly, we also report that circulating Spike protein can be linked to EVs in the absence of viral RNA in the vesicles. These results suggest that the persistence of Spike protein and/or viral RNA could be contributing factors for the development of PASC.

The mechanisms of how SARS-CoV-2 can persist and cause prolonged and various symptoms long after acute infection has yet to be elucidated. Several theories have been proposed, including that parts of the SARS-CoV-2 genome can be incorporated into the DNA of infected cells and that fragments of the SARS-CoV-2 genome may be transcribed at random, but the evidence for this is disputed^18, 19^. Alternative explanations for SARS-CoV-2 persistence include mechanisms that are used by other RNA viruses to persist in host cells, including evasion of the host immune response, infection of long-lived cells, and/or infection of immune privileged tissues such as the brain, eyes, or testes, or diminished cell-mediated immunity within a patient^2, 20, 21^. Additionally, SARS-CoV-2 may be able to persist due to an insufficient immune response during acute COVID-19^22, 23^.

Our results revealed that most patients did not have both Spike protein and viral RNA in plasma simultaneously, suggesting that the persistence of viral particles is probably not due to actively replicating virus. Replication of coronaviruses produces a set of sub-genomic RNA molecules with heterogeneous 5’ ends but identical 3’ ends. All genomic and sub-genomic SARS-CoV-2 RNAs contains the nucleocapsid (N) gene sequence due to its position at the 3’ end of the viral genome^24, 25^. For this reason, N represents the most abundant and the most suitable target for SARS-CoV-2 RNA detection. We were able to detect low copies of N gene in plasma samples of approximately 35% of the acute COVID-19 group and 55% of PASC+ve patients by highly sensitive ddPCR method which normally would have been undetectable by conventional RT-PCR^26^. While we didn’t perform whole RNA sequencing, it is possible that we may be detecting viral RNA fragments as we found a decrease in viral RNA in most of the samples after RNAse treatment. Some PASC-ve individuals had detectable levels of Spike protein or viral RNA in plasma, and one may speculate SARS-CoV-2 to be more immunogenic in these patients which may prevent PASC. Alternatively, some patients with PASC had neither Spike protein nor viral RNA detected in plasma. In these patients, PASC could be possibly attributed to prolonged immune dysregulation, microthrombosis after acute COVID-19 or potentially worsening of pre-existing conditions.

Similar to previous reports that SARS-CoV-2 viremia is positively correlated with acute COVID-19 severity and is a predictor of mortality^27, 28^, we found that Spike protein is positively correlated with peak WHO score (p=0.0351) during hospitalization. We also observed that Spike protein is positively correlated with length of hospitalization (p=0.0359) which is of particular concern because prior research has shown that longer duration of SARS-CoV-2 viremia is associated with greater risk of mortality^29^. It also appears that a decreased production of early functional, neutralizing antibodies is a predictor of COVID-19 severity and may also explain why Spike protein is correlated with peak WHO score and length of hospitalization due to lack of clearance by cell-mediated immunity^22, 23^. However, we did not observe any correlation of viral RNA with either disease severity or length of hospitalization in patients with acute disease.

It is now widely recognized that endothelial dysfunction and micro-thromboses play a significant role in acute infection and we see further evidence of this with our finding that D-dimer is positively correlated with circulating levels of total plasma Spike protein (p=0.0249) and EV-associated Spike protein (p<0.01) across all three groups of acute COVID-19 severity. D-dimer is a marker of coagulopathy^30, 31^ and is highly correlated with the COVID-19 disease severity^32, 33^. Park et al^32^ recently reported that D-Dimer can also form immune complexes with SARS-CoV-2 Spike protein and bound antibodies resulting in activation of monocytes to produce proinflammatory cytokines. Moreover, endothelial dysfunction may be caused by both direct infection of endothelial cells^34^ and by the SARS-CoV-2 components in the bloodstream^35^. Perico et al^35^ studied the effects of SARS-CoV-2-derived Spike protein alone on HMVECs and found that treatment of these cells with Spike protein led to dose-dependent increases in ICAM-1 and von Willebrand Factor deposition, as well as increased deposition of complement protein C3 on the surface of HMVEC when cells were treated with higher concentrations of Spike protein^35^. It is intriguing that we showed increased levels of Spike protein in PASC+ve patients, indicating that Spike protein-driven endothelial activation may persist and perhaps contribute to increased risk of clotting and endothelial dysfunction during PASC.

Immune system dysregulation and a persistent proinflammatory state may also contribute to the development of PASC. Elevated markers of inflammation, including IL-6 and C-reactive protein, remain elevated in some patients who exhibit PASC symptoms^36, 37^. Our findings of increased copies of viral RNA in the PASC+ve group suggest that, along with Spike protein, may also be contributing to persistent inflammation. It has been shown previously that circulating viral RNA could contribute to inflammation by activation of sensory Toll-like receptors (TLR), including TLR-3 and TLR-7^38^. Viral RNA can stimulate TLR3 and activate the innate immune system to produce inflammatory cytokines, including IL-6, or can stimulate TLR7 and induce the production of interferons^38^.

In our recently published study, we found that levels of EN-RAGE, IL-18R1, and tissue factor (TF) in EVs were especially correlated with COVID-19 severity and length of hospitalization. We also found that EVs from hospitalized COVID-19 patients induce apoptosis of endothelial cells *in vitro*^10^. Here, we observed that EVs carry Spike protein in both acute COVID-19 and PASC patients. Given that SARS-CoV-2 replicates in cytosol using endosomal pathway^39^ and we observed Spike protein associated with EVs, it may be possible for SARS-CoV-2 to usurp EV biogenesis pathways and conceal itself within EVs as protection against neutralizing antibodies. However, we did not observe viral RNA in EVs in post-COVID patients, though it is possible that there may be very low copy number present below the limit of detection of ddPCR. Removal of surface proteoglycans from EVs with Heparinase II or treatment with Proteinase K showed a decrease in Spike levels suggesting Spike protein to be partly present on the surface of EVs via heparin sulfate interactions. Numerous studies shed light on the key role of Heparan sulfate proteoglycans with the SARS-CoV-2 binding at the cell membrane. Heparan sulfate interacts with the adjacent residues of the ACE2-binding site at the receptor binding domain (RBD) of the S1 subunit of the SARS-CoV-2 trimeric S-protein^40, 41^.

Finally, we must acknowledge the limitations of our study. PASC, thus far, has been very difficult to study due to the heterogeneity of the clinical presentations and the lack of a uniform definition. We defined our PASC positive patients as those with any self-reported symptoms via survey or to a provider. Self-reported symptoms and surveys by their very nature are subjective and likely result in some error in designating patients into a defined group. Many of the included patients also had comorbidities such as underlying cardiovascular disease, obesity, or diabetes, again making it difficult to ascertain if reported symptoms are worsening underlying chronic conditions or from past COVID infection. Another limitation is that our acutely infected cohort was all patients that needed to be hospitalized for their illness while the patients in PASC cohort also included individuals that had mild initial infection. We also had a limited sample size of patients and future studies using a larger cohort are warranted. In conclusion our study reveals intriguing findings that the persistence of Spike protein and viral RNA fragments from SARS-CoV-2 is associated with PASC and may contribute to its development.

## Data Availability

All data produced in the present work are contained in the manuscript

## ACKNOWLEDGEMENTS

We acknowledge Ling Chen, Department of Internal Medicine, KUMC, for her help in processing blood specimens. We also acknowledge the significant amount of time and effort Ashok Kumar has devoted to Nanosight Tracking Analyses. We would also like to thank Luigi Boccardi and Maggie Chen for their assistance in maintaining clinical data and COVID-19 biorepository.

