## Supplemental Table and Figure for "Persistent Presence of Spike protein and Viral RNA in the Circulation of Individuals with Post-Acute Sequelae of COVID-19"

**Supplementary Table 1: Correlation Analyses of Clinical Characteristics with Viral RNA, Spike level in Plasma and Plasma-derived EVs in COVID-19 Individuals**

| | **Viral RNA [Correlation (p-value)]** | **Plasma spike [Correlation (p-value)]** | **SEVs spike [Correlation (p-value)]** |
|---|---|---|---|
| **Acutely -infected COVID-19 Cohort** | | | |
| *Age* | 0.0981 (0.2948) | 0.0398 (0.6728) | -0.0061 (0.9597) |
| *BMI* | 0.04 (0.6695) | 0.0809 (0.3898) | -0.2108 (0.0776) |
| *Peak WHO Score* | 0.0497 (0.6026) | **0.2002 (0.0351) *** | 0.0716 (0.5646) |
| *Length of hospitalization* | 0.1379 (0.2549) | **0.253 (0.0359) *** | 0.1579 (0.324) |
| *WBC Count* | 0.1009 (0.2856) | 0.0567 (0.5508) | 0.1446 (0.2325) |
| *Lymphocyte Count* | -0.133 (0.1806) | -0.0676 (0.4995) | -0.2072 (0.1031) |
| *Creatinine* | 0.1477 (0.1153) | 0.0055 (0.9536) | 0.0509 (0.6758) |
| *Lactate Dehydrogenase* | **0.2532 (0.0273) *** | 0.1904 (0.0994) | 0.1781 (0.2309) |
| *Ferritin* | 0.1543 (0.1586) | 0.0627 (0.5689) | 0.1888 (0.1892) |
| *D.Dimer* | 0.0863 (0.4136) | **0.2338 (0.0249) *** | **0.3544 (0.0092) **** |
| *C-Reactive Protein* | 0.1335 (0.2149) | 0.1451 (0.1774) | -0.1266 (0.3713) |
| **Post-Acute Infection Cohort** | | | |
| *Age* | 0.0456 (0.761) | -0.0231 (0.8774) | -0.1102 (0.6343) |
| *Days between last positive test and study enrollment* | -0.1712 (0.2552) | 0.1185 (0.4326) | -0.1635 (0.4789) |
| *BMI* | 0.0717 (0.6646) | -0.1851 (0.2594) | -0.1465 (0.5377) |
| *Length of Hospitalization* | **0.3338 (0.0435)*** | -0.1634 (0.3339) | 0.442 (0.0581) |

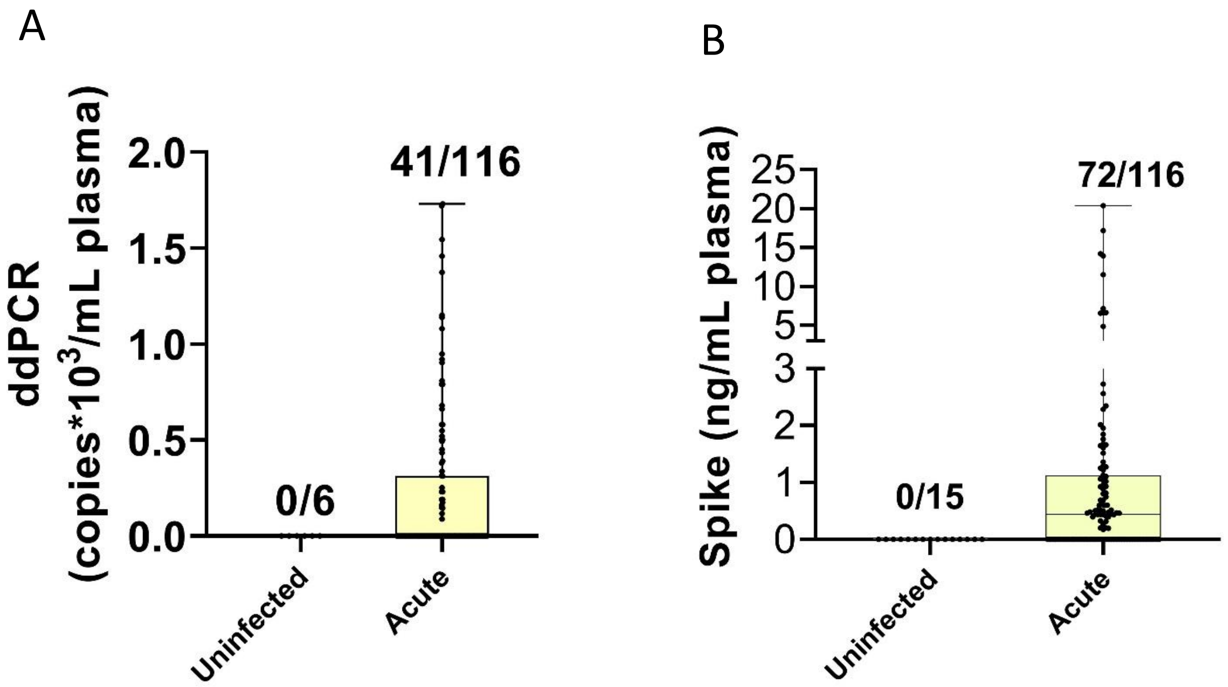


**Supplementary figure 1:** Analysis of SARS-CoV-2 RNA (A) and Spike protein levels (B) in the plasma from *Acute* COVID-19 patients and never infected with COVID-19 controls as measured by dd-PCR and ELISA.
